# Muscle stability deficits and musculoskeletal complaints in football players: Adaptive Force Ratio outperforms conventional strength parameters—a cross-sectional study with preliminary follow-up

**DOI:** 10.64898/2026.07.07.26357205

**Authors:** Laura V Schaefer, Frank N Bittmann, Jaali Ulrich, Robert Prill, Roland Becker

## Abstract

Strength-based screening has documented limitations. Adaptive Force (AF) offers an alternative—it captures the adaptive holding capacity under rising external load, not addressed by strength testing. This study examined the association between AF-based muscle stability and musculoskeletal complaints in 23 football (soccer) players, comparing AF-derived and conventional strength parameters, with preliminary follow-up. AF and maximal voluntary isometric contraction (MVIC) were measured across five bilateral muscle groups (knee extensors/flexors; hip flexors/adductors/abductors). AF parameters (maximal isometric AF, maximal AF, AF-Ratio), MVIC and hamstrings-to-quadriceps (H:Q) ratio were compared by complaint status assessed via questionnaire at baseline and six-month follow-up (n=13).

Stability deficits were strongly associated with complaints (OR=54.0, 82% side concordance). AF-Ratio discriminated between players with and without complaints (*d*=−1.47, AUC=0.93), strongest for hip abductors (*d*=−1.64). Players with follow-up complaints showed lower baseline AF-Ratio (*d*=−1.45, AUC=0.86) and more stability deficits (*d*=1.67). MVIC and H:Q ratio discriminated in neither analysis (*p*>0.500, AUC≤0.44); AF-Ratio outperformed both at baseline (*p*≤0.002) and follow-up (*p*≤0.028).

AF-based stability assessment may capture neuromuscular deficits related to complaints that conventional strength testing misses, with preliminary follow-up data suggestive of predictive value. The concept of functional instability syndrome provides a mechanistic framework for non-contact injuries and complaints. AF-based stability assessment may hold potential for screening and return-to-sport decisions—pending prospective validation.

## 1 Background

Injuries remain a persistent challenge in football (soccer); the lower limbs are predominantly affected, and most injuries occur in non-contact situations.[1,2] Although prevention programs and strength screening are widely adopted, incidence rates have increased or remain high.[1–4] Valid screening parameters for identifying players at risk are still lacking: conventional strength-based testing has shown limited predictive value,[5–10] also for re-injury [11]—in a meta-analysis, none of 13 strength-related variables was associated with hamstring injury risk.[5] The strongest predictor of injury is previous injury.[5,12] Where strength has been associated with pain, effects are at most moderate and rest on low-certainty evidence, with findings varying across muscle groups and testing procedures.[13–15] This cumulative evidence suggests that strength-based methods may have reached their limits, warranting exploration of whether a different dimension of neuromuscular function may be more relevant to musculoskeletal complaints and injuries.

Muscle stability assessed by Adaptive Force (AF) offers an alternative: in contrast to conventional strength tests (pushing/pulling actions), it captures a motor task that more closely reflects injury-prone movements, which involve active deceleration and dynamic stabilization.[1,4,16,17] AF assesses the neuromuscular capacity to maintain position against increasing external loads during holding isometric muscle action (HIMA).[18–26] If the muscle cannot sustain position, involuntarily yielding to lengthening at submaximal levels, it is classified as unstable. In a companion study,[27] stability deficits were highly prevalent in semi-professional football players (83% of players, 31% of muscle groups), with a selective impairment of holding capacity (∼61% lower than maximal force output) while maximal pushing strength (maximal voluntary isometric contraction; MVIC) remained unaffected. The ratio of holding capacity to peak force (AF-Ratio) emerged as a muscle-independent stability parameter.[18–24,27] Previous studies revealed that the holding capacity responds instantaneously to various impairing stimuli (e.g., disturbed proprioception, nociception, negative emotions, post-infectious conditions) with reductions of 39–53%,[18–24] suggesting that muscle stability reflects a broader dimension of neuromuscular integrity beyond physical conditioning. Its association with musculoskeletal complaints in athletes has not been investigated. Specifically in sports where dynamic stabilization and active deceleration are critical, AF may provide novel insight into injury development.

The study’s primary aim was to examine the association between muscle stability deficits of the lower extremities and musculoskeletal complaints in semi-professional football players. The secondary aim was to compare the discriminative capacity of AF-Ratio with conventional strength parameters (MVIC, hamstrings-to-quadriceps (H:Q) ratio) for distinguishing players with and without complaints. It was hypothesized that muscle stability deficits would be associated with musculoskeletal complaints, in contrast to conventional strength parameters. As a preliminary prospective element, a six-month follow-up examined whether baseline stability deficits were associated with subsequent injuries or complaints.

## 2 Methods

This cross-sectional study with preliminary follow-up analyzed the same dataset as Schaefer et al.[27] Measurements were performed during the preseason (July/August 2025).

### 2.1 Participants

Twenty-three male semi-professional football players participated (23.7 ± 5.7 years, 184.4 ± 5.6 cm, 79.3 ± 6.9 kg, training: 6.3 ± 1.0 h/week). Inclusion criterion: active participation in training (≥3 sessions/week). Exclusion criteria: acute injury preventing testing, neurological conditions affecting motor control. Sample size was determined by team availability; based on previous AF effect sizes (*d* = 2.72),[18] 23 players were considered adequate.

This study was conducted in accordance with the Declaration of Helsinki and approved by the Ethics Committee of University of Potsdam (protocol number: 35/2018; 17 October 2018). All participants provided written informed consent.

### 2.2 Assessment of musculoskeletal complaints

A structured questionnaire recorded any musculoskeletal complaints, including injuries, irrespective of time-loss or medical attention: current complaints (pain intensity 0–10), those within the past three months, and those beyond three months (each with region, affected side, injury mechanism, and date). A six-month follow-up survey (January 2026) recorded subsequent complaints/injuries without reassessing force parameters.

### 2.3 Force assessment and stability classification

Five muscle groups were tested bilaterally (randomized starting side; Supplementary Methods, Supplementary Figure S1): knee extensors (MQF) and flexors (HAM), hip flexors (HFL), adductors (ADD) and abductors (ABD). AF was assessed by objectified, standardized manual muscle tests (3 trials per muscle and side) using a dedicated handheld device incorporating strain gauges (precision: 1.0 ± 0.1%) and kinematic sensors (BNO055, Bosch, Germany) for simultaneous recording of force and angular velocity (sampling rate: 180 Hz). The MVIC (2 trials per muscle and side) was recorded with the same device, except for MQF, measured by handheld dynamometers (MicroFET2/MicroFET3, Hoggan Scientific, USA). Each participant was assessed by one of two experienced examiners throughout. Test-retest reliability of force application within and between examiners was demonstrated previously (intraclass correlation coefficient, ICC(3,1) = 0.989).[28]

AF differs from MVIC in requiring continuous adaptation to an increasing external force applied by the examiner to maintain position (adaptive HIMA), whereas MVIC involves pushing against fixed resistance without adaptive demands (pushing isometric muscle action; PIMA).

The following AF parameters were extracted from the same trial (Fig. 1; detailed in Supplementary Methods): maximal isometric AF (AFiso_max_, holding capacity; highest force under quasi-static conditions), peak AF (AF_max_), and AF-Ratio (AFiso_max_/AF_max_). From MVIC tests, peak force and H:Q ratio (HAM/MQF) were obtained. Force values (N) were converted to torque (Nm) using the individual lever arm; AF-Ratio (within-trial ratio) is independent of this conversion.

**Fig. 1.**
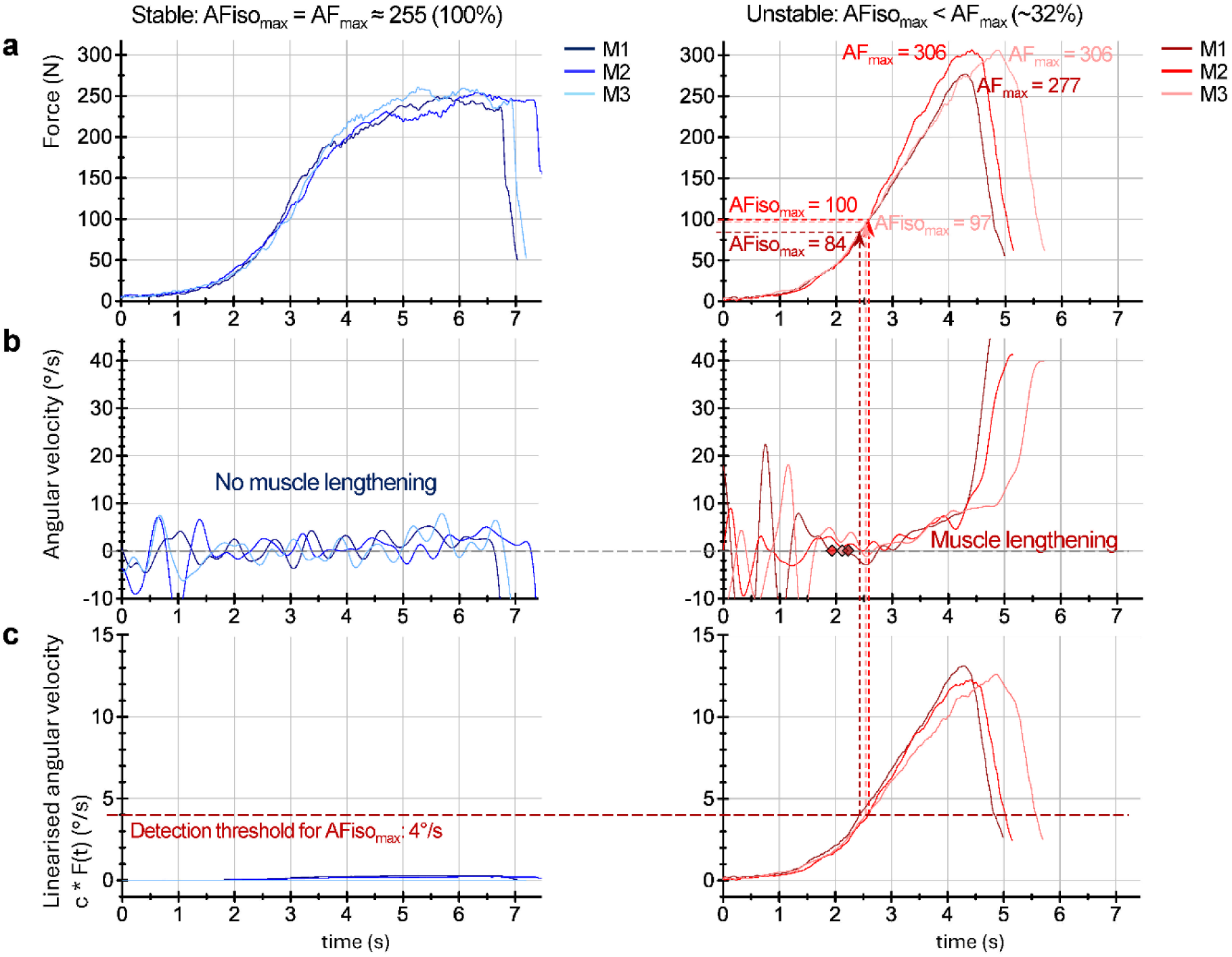
Exemplary Adaptive Force measurement curves for a stable and an unstable muscle. Force (N; **a**), angular velocity (°/s; **b**) and linearized angular velocity (°/s; **c**) for three consecutive AF trials (M1–M3) of hip flexors from two players. Left panels: stable muscle—angular velocity oscillates around zero throughout (**b**), and linearized angular velocity (**c**) remains below the detection threshold for AFiso_max_ (horizontal dashed line), indicating that the starting position was maintained quasi-static until peak force; AFiso_max_ = AF_max_ (100%) (Player A, right HFL, MVIC 262 N; female examiner). Right panels: unstable muscle—diamond markers in (**b**) indicate zero-crossings representing the onset of muscle lengthening despite further force increase (**a**). The linearized angular velocity (**c**) exceeds the detection threshold during force increase, marking AFiso_max_ (vertical dashed lines); AFiso_max_ is substantially lower than AF_max_ (∼32%) (Player B, left HFL, MVIC 284 N; female examiner). Angular velocity (**b**): gyroscope signal (gX axis), low-pass filtered (Butterworth, 2 Hz, 4^th^ order); dashed horizontal line indicates zero. Linearized angular velocity (**c**): c · F(t), where c is the slope of the least-squares fit (Supplementary Methods). In the absence of muscle lengthening, c approximates zero and the linearized angular velocity remains near zero despite increasing force. AF = Adaptive Force, AFisomax = maximal isometric AF, AFmax = maximal AF, HFL = hip flexors, MVIC = maximal voluntary isometric contraction.

For statistics, the maximum of the two MVIC trials and the average of the three AF trials were used, yielding one value per parameter at muscle group level (n = 230: 5 muscle groups × 2 sides × 23 players). Stability was classified from AF trials using a gyroscope-based algorithm validated against the examiners’ ratings (stable/unstable/borderline; accuracy: 98.5%, weighted κ = 0.96; Supplementary Methods) and aggregated across trials per muscle group.

### 2.4 Statistical analysis

Descriptive statistics are presented as arithmetic mean and standard deviation (M ± SD). For pooled absolute parameters (MVIC, AF_max_, AFiso_max_), values were z-standardized within each muscle group before averaging across muscles and sides, to account for between-muscle differences in absolute torque. Similar force application across stability categories was confirmed by non-significant differences in the rate of force development (z-standardized, *p* = 0.514; Supplementary Data).

#### Complaint association (player level, n = 23)

A stability deficit was defined as ≥1 non-stable classification (unstable/borderline) at aggregated level. Players were ‘symptomatic’ if they reported at least one complaint (current/recurrent within past 12 months/non-contact injuries within past 6 months; irrespective of body region). The association between stability status (completely stable vs. stability deficit) and complaint status (complaints vs. complaint-free) was analyzed by Fisher’s exact test for all complaints and lower-extremity complaints only; odds ratio (OR), sensitivity, specificity, positive predictive value and negative predictive value are reported. Force parameters (averaged across both sides per player) were compared between complaint groups using Welch’s t-tests, pooled and per muscle group. For direct comparability with H:Q ratio, AF-Ratio was additionally averaged across HAM and MQF only. Correlation between H:Q ratio and AF-Ratio was assessed using Spearman’s rank correlation ρ. The numbers of stability deficits and unstable muscles were compared using Mann-Whitney U tests.

Side concordance was examined descriptively in players with lower-extremity complaints, defined as more deficits on the complaint side (unilateral) or on both sides (bilateral). Where a specific complaint side was identifiable, AF-Ratio was compared between sides using paired t-test (two-tailed). Associations between muscle groups and complaint regions were reported descriptively owing to the small sample size and anatomical overlap.

#### Preliminary follow-up

Baseline AF-Ratio, MVIC, and H:Q ratio were compared between players with and without follow-up complaints using Welch’s t-tests, and the number of stability deficits using the Mann-Whitney U test.

Discriminative capacity for complaint status was assessed at player level for AF-Ratio, AFiso_max_, AF_max_, MVIC, and H:Q ratio as well as for the number of stability deficits using receiver operating characteristic (ROC) analysis.[29] As conventional strength-based screening assumes that lower values indicate higher risk, all classifiers were defined a priori in this direction, except for the number of stability deficits (higher values). An area under the curve (AUC) below 0.5 indicates that a parameter does not discriminate in the hypothesized direction. AUCs with logit-transformed 95% confidence intervals (CI) were compared pairwise against AF-Ratio by DeLong’s test for correlated AUCs,[29] both at baseline and follow-up. Given the small complaint-free group, a stratified bootstrap (10,000 resamples) of the difference in AUC was performed as confirmation.

Effect size was expressed as Cohen’s *d* [0.2 (small), 0.5 (medium), 0.8 (large)].[30] Significance was set at *p* < 0.05. Analyses were performed using Python 3.12 and jamovi 2.7.18; the large language model Claude (Anthropic) assisted with data processing scripts.

## 3 Results

### 3.1 Association between musculoskeletal complaints and stability deficits

Nineteen of 23 players (83%) reported complaints within the past year, four (17%) were complaint-free. 84% of symptomatic players reported a history of complaints/injuries extending beyond the past year (i.e. recurrent), whereas only one complaint-free player did. Lower-extremity complaints were present in 17 of 19 symptomatic players, 2 of 19 reported upper-extremity complaints only (both with stability deficits). Eighteen of 19 symptomatic players had at least one stability deficit at aggregated level, 3 of 4 complaint-free players were completely stable.

Complaints and stability deficits were strongly associated (OR = 54.0), including when restricted to lower-extremity complaints (Table 1). Symptomatic players had more stability deficits (4.6 ± 2.5 vs. 0.8 ± 1.5; *p* = 0.011, *d* = 1.63) and more unstable muscles (3.6 ± 2.5 vs. 0.5 ± 1.0; *p* = 0.019, *d* = 1.31) than complaint-free players.

**Table 1.** Association between stability deficits and musculoskeletal complaints.

| Level | Group | deficits | no deficits | OR (95%-CI) | $p$ | Sens | Spec | PPV | NPV |
| --- | --- | --- | --- | --- | --- | --- | --- | --- | --- |
| All complaints | Complaints (n = 19) | 18 | 1 | 54.0<br>(2.6–1117) | 0.009 | 95% | 75% | 95% | 75% |
|  | Complaint-free (n = 4) | 1 | 3 |  |  |  |  |  |  |
| LE complaints | Complaints (n = 17) | 16 | 1 | 16.0<br>(1.2–211) | 0.040 | 94% | 50% | 84% | 75% |
|  | Complaint-free (n = 6) | 3 | 3 |  |  |  |  |  |  |

Stability deficit defined as $\geq 1$ non-stable classification at aggregated level. Values of Fisher's exact test compared between complaint vs. complaint-free players are given.
LE = lower extremity; NPV = negative predictive value; PPV = positive predictive value; Sens = sensitivity, Spec = specificity.

Stability deficits were detected in at least one muscle in 83–100% of players across all complaint regions (Fig. 2a, Supplementary Table S2). ABD showed the most consistent association (67–100%; mean: 89%), while MQF the lowest (0–67%, mean: 26%), largely confined to load-related complaints; correspondingly, 80% of MQF-unstable players reported load-related complaints (Fig. 2b). For the remaining muscles, deficits frequently occurred without corresponding regional complaints (0–35%).

**Fig. 2.**
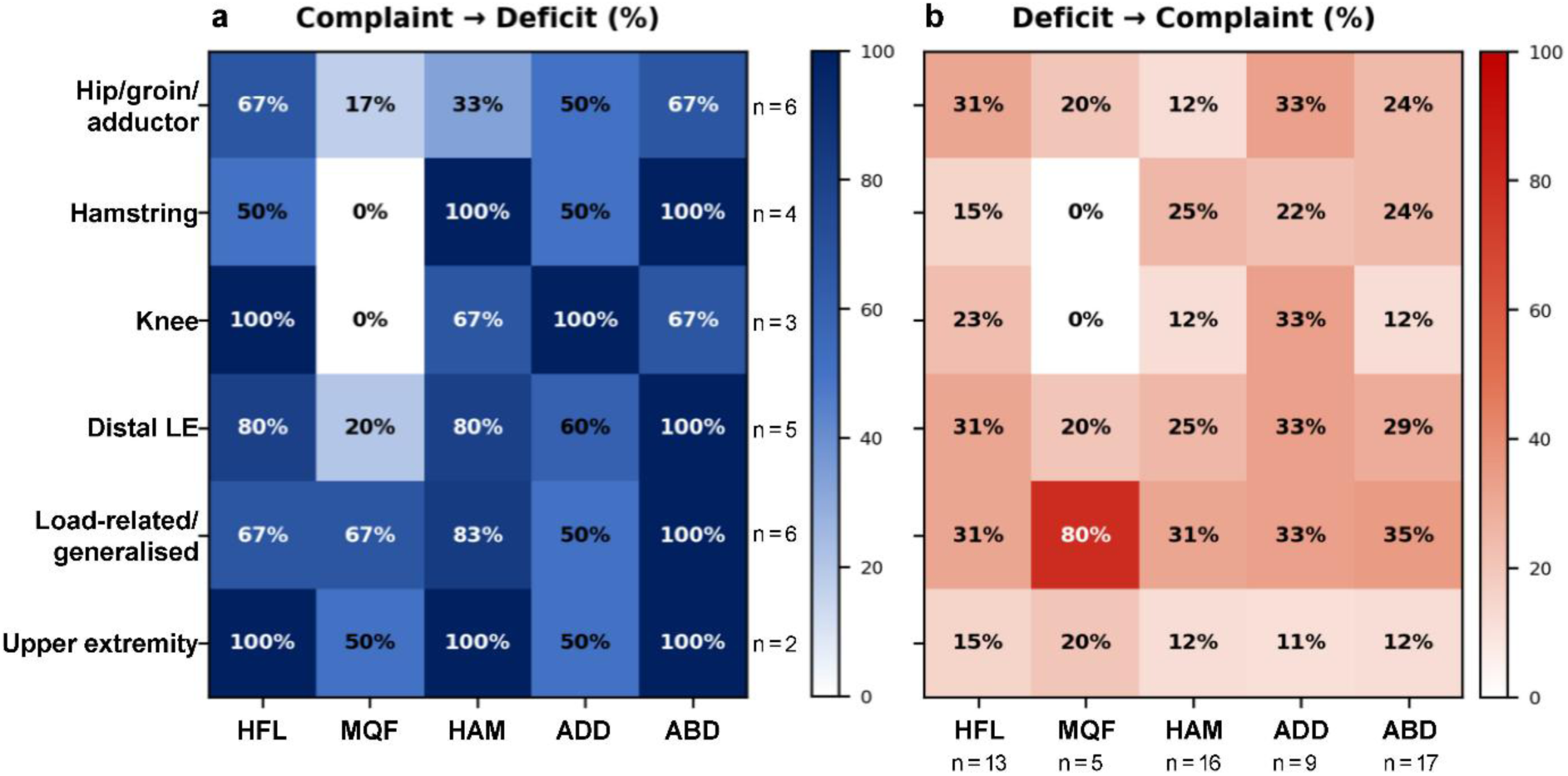
Associations between complaint region and stability deficits per muscle group. (a) : Complaint → Deficit: proportion (%) of players with complaints in the given region who had stability deficits; n = number of players with complaints in that region. (b) : Deficit → Complaint: proportion (%) of all players with a stability deficit in the given muscle group who reported complaints in that region; n = number of players with a deficit in that muscle group. Players with complaints in multiple regions are counted in each applicable row. ABD = hip abductors, ADD = hip adductors, HAM = knee flexors (hamstrings), HFL = hip flexors, LE = lower extremity, MQF = knee extensors (quadriceps femoris). Load-related/generalized: acute muscle soreness or pain related to training load.

Between players with and without complaints (Fig. 3a, Table 2), MVIC and AF_max_ did not differ significantly, AFiso_max_ presented a large but non-significant effect (*d* = −1.00) and AF-Ratio showed the clearest separation (*d* = −1.47), with the largest effect for ABD. ABD instability on at least one side was present in 79% of symptomatic players but in none of the complaint-free players, followed by HAM (68% vs. 25%), HFL (47% vs. 0%), ADD (32% vs. 0%) and MQF (21% vs. 0%). The H:Q ratio did not differ significantly between players with and without complaints (*d* = 0.18), whereas the AF-Ratio restricted to HAM and MQF did (*d* = −0.97). The two ratios were uncorrelated (*ρ* = −0.18, *p* = 0.421).

**Fig. 3.**
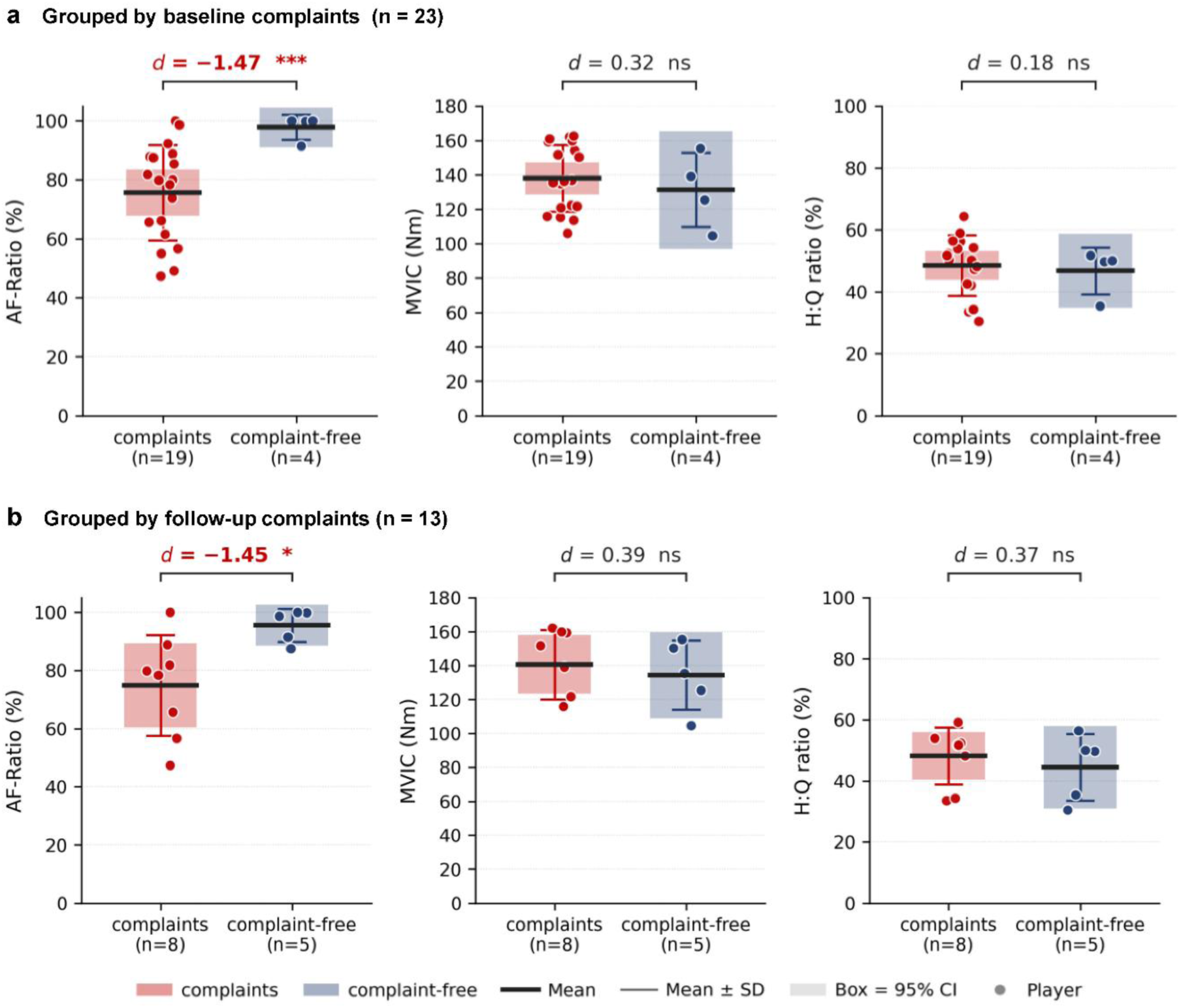
Comparison of baseline force parameters grouped by complaint status at baseline and at follow-up. Plots show individual player values (dots) with the group mean (thick line), mean ± SD (whiskers) and 95% CI (shaded box) of AF-Ratio (%), MVIC (Nm), and H:Q ratio (%). **(a)** Cross-sectional comparison at baseline: players with complaints (n = 19) vs. complaint-free (n = 4). **(b)** Prospective comparison based on baseline parameters: players with subsequent complaints within the 6-month follow-up (n = 8) vs. complaint-free (n = 5). AF-Ratio and MVIC are pooled across all muscle groups; for MVIC (Nm), *d* and *p* were computed on values z-standardized within each muscle to account for differences between muscle groups. As AF-Ratio is bounded at 100%, the symmetric SD whiskers and 95% CI of the complaint-free group extend slightly above this ceiling; values >100% are not physiologically meaningful. Cohen’s d is positive when complaints > complaint-free; *p*-values from Welch’s t-test (two-sided). \**p* < 0.05; \*\*\**p* < 0.001; ns = not significant. AF-Ratio = max. isometric Adaptive Force (AF) / max. AF; MVIC = max. voluntary isometric contraction; H:Q ratio = MVIC hamstrings / MVIC quadriceps.

**Table 2.** Comparison of force parameters in players with and without complaints. Force parameters are given as player means (M ± SD) pooled across all five muscle groups (H:Q as the HAM/MQF ratio) and both sides. Players with (n = 19) and without (n = 4) complaints were compared at baseline (cross-sectional); baseline force values were additionally used for a preliminary prospective comparison of players with (n = 8) and without (n = 5) subsequent complaints at follow-up. For cross-sectional comparison AF-Ratio is additionally shown per muscle group. Mean differences (players with − without complaints) with 95% confidence interval [CI] from Welch’s t-test are shown. For the absolute parameters pooled across muscles, p-values and Cohen’s d are based on z-standardized values to account for differences between muscle groups, whereas the mean differences and CIs are reported in the original units.

| Parameter | Muscle | Complaints | Complaint-free | Mean difference [CI] | p | d |
| --- | --- | --- | --- | --- | --- | --- |
| **Baseline** |  |  |  |  |  |  |
| **MVIC (Nm)** | pooled | 137.94 ± 19.30 | 131.19 ± 21.59 | +6.75 [-25.41, +38.91] | 0.594 | +0.32 |
| **H:Q ratio (%)** | pooled | 48.50 ± 9.79 | 46.76 ± 7.55 | +1.74 [-9.31, +12.79] | 0.707 | +0.18 |
| **AF<sub>max</sub> (Nm)** | pooled | 140.87 ± 14.65 | 135.36 ± 22.69 | +5.51 [-29.07, +40.09] | 0.589 | +0.44 |
| **AFiso<sub>max</sub> (Nm)** | pooled | 106.56 ± 24.65 | 132.67 ± 19.83 | -26.11 [-55.15, +2.93] | 0.093 | -1.00 |
| **AF-Ratio (%)** | pooled | 75.57 ± 16.22 | 97.81 ± 4.25 | -22.24 [-31.20, -13.28] | **<0.001** | -1.47 |
| | HFL | 71.51 ± 28.46 | 100.00 ± 0.00 | -28.49 [-42.21, -14.77]† | **<0.001** | -1.08 |
| | MQF | 91.19 ± 17.99 | 100.00 ± 0.00 | -8.81 [-17.48, -0.14]† | **0.047** | -0.53 |
| | HAM | 67.34 ± 27.01 | 91.33 ± 17.35 | -23.99 [-49.55, +1.57] | 0.062 | -0.93 |
| | ADD | 88.33 ± 16.20 | 100.00 ± 0.00 | -11.67 [-19.48, -3.86]† | **0.006** | -0.78 |
| | ABD | 59.47 ± 25.18 | 97.71 ± 3.93 | -38.24 [-50.94, -25.54] | **<0.001** | -1.64 |
| | HAM+MQF | 79.27 ± 17.99 | 95.66 ± 8.67 | -16.39 [-29.80, -2.98] | **0.022** | -0.97 |
| **Follow-up** |  |  |  |  |  |  |
| **MVIC (Nm)** | pooled | 140.67 ± 20.46 | 134.26 ± 20.44 | +6.41 [-20.13, +32.94] | 0.502 | +0.39 |
| **H:Q ratio (%)** | pooled | 48.17 ± 9.31 | 44.47 ± 10.94 | +3.70 [-10.04, +17.44] | 0.549 | +0.37 |
| **AF<sub>max</sub> (Nm)** | pooled | 143.53 ± 17.30 | 129.93 ± 17.33 | +13.60 [-8.89, +36.08] | 0.174 | +0.86 |
| **AFiso<sub>max</sub> (Nm)** | pooled | 108.19 ± 33.43 | 123.86 ± 15.63 | -15.67 [-46.07, +14.72] | 0.359 | -0.47 |
| **AF-Ratio (%)** | pooled | 74.83 ± 17.29 | 95.49 ± 5.62 | -20.66 [-35.59, -5.74] | **0.012** | -1.45 |

† For HFL, MQF and ADD, all complaint-free players scored 100% ( $SD = 0$ ); the CI therefore reflects the variance of the symptomatic group only.
Significant results are displayed in bold.
ABD = hip abductors; ADD = hip adductors; AFiso<sub>max</sub> = max. isometric Adaptive Force; AF<sub>max</sub> = max. AF; AF-Ratio = AFiso<sub>max</sub> / AF<sub>max</sub>; H:Q ratio = MVIC hamstrings / MVIC quadriceps; HAM = hamstrings; HFL = hip flexors; MQF = quadriceps femoris; MVIC = max. voluntary isometric contraction.

In 14 of 17 players with lower-extremity complaints (82%), more stability deficits were found on the complaint side—or on both sides for bilateral complaints (side concordance). Two players showed more deficits contralaterally, one had no deficits. AF-Ratio (pooled across muscles) was significantly lower on the complaint vs. complaint-free side (n = 10; 76.2 ± 20.8% vs. 89.4 ± 14.7%; *p* = 0.038, *d* = −0.77).

Beyond the group-level differences, the discriminative capacity confirmed the pattern. AF-Ratio separated players with and without complaints with an AUC of 0.93, significantly outperforming MVIC (AUC = 0.41), H:Q ratio (AUC = 0.43) and AF_max_ (AUC = 0.40; Table 3). AUC values below 0.5 indicate higher parameter values in symptomatic than in asymptomatic players (i.e. discrimination opposite to that hypothesized). AFiso_max_ (AUC = 0.79) and the number of stability deficits (AUC = 0.91) discriminated comparably to AF-Ratio (Table 3).

**Table 3.** Discriminative capacity for baseline and follow-up complaints using receiver operating characteristic (ROC) analysis.

| Parameter | AUC | 95% CI | ΔAUC | *p* |
| --- | --- | --- | --- | --- |
| **Baseline** |  |  |  |  |
| **AF-Ratio (%)** | **0.934** | **0.727–0.987** | **—** | **—** |
| MVIC (Nm) | 0.408 | 0.164–0.707 | +0.526 | 0.002 |
| H:Q ratio (%) | 0.434 | 0.219–0.678 | +0.500 | <0.001 |
| AF<sub>max</sub> (Nm) | 0.395 | 0.088–0.815 | +0.539 | 0.036 |
| AFiso<sub>max</sub> (Nm) | 0.789 | 0.434–0.948 | +0.145 | 0.334 |
| Stability deficits (n) | 0.914 | 0.650–0.984 | +0.020 | 0.677 |
| **Follow-up** |  |  |  |  |
| **AF-Ratio (%)** | **0.863** | **0.481–0.977** | **—** | **—** |
| MVIC (Nm) | 0.350 | 0.113–0.695 | +0.513 | 0.014 |
| H:Q ratio (%) | 0.375 | 0.118–0.730 | +0.488 | 0.028 |
| AF<sub>max</sub> (Nm) | 0.225 | 0.048–0.626 | +0.638 | 0.001 |
| AFiso<sub>max</sub> (Nm) | 0.600 | 0.269–0.859 | +0.263 | 0.065 |
| Stability deficits (n) | 0.900 | 0.558–0.985 | –0.037 | 0.364 |

$\Delta$ AUC = AUC(AF-Ratio) – AUC(parameter); *p*-values from DeLong's test for correlated AUCs.
Classifier direction specified a priori as lower value for complaints, except for the number of stability deficits (higher value indicating complaints).
AFiso<sub>max</sub> = max. isometric Adaptive Force; AF<sub>max</sub> = max. AF; AF-Ratio = AFiso<sub>max</sub> / AF<sub>max</sub>; AUC = area under the curve;
H:Q ratio = MVIC hamstrings / MVIC quadriceps; MVIC = max. voluntary isometric contraction.

### 3.2 Preliminary Follow-up: stability deficits and prospective complaints

Follow-up data were available for 15 players (65%); 13 were included after excluding two whose only baseline complaint was training-related, likely reflecting transient stability deficits—neither reported follow-up complaints. Three of the four fully stable players reported no subsequent complaint/injury, one had a knee injury without time-loss. Seven of nine players with stability deficits had subsequent complaints/injuries (5 non-contact, 2 contact, 6 with time-loss); two players stayed complaint-free.

Players with subsequent complaints at follow-up had significantly lower baseline AF-Ratio than those without (*d* = −1.45) and more stability deficits (4.6 ± 2.5 vs. 1.0 ± 1.4; *p* = 0.021, *d* = 1.67); baseline MVIC and H:Q ratio differed non-significantly between both groups (Fig. 3b, Table 2). Baseline AF-Ratio also significantly outperformed MVIC and H:Q ratio in discriminating players with subsequent complaints (AUC = 0.86 vs. 0.35 and 0.38; Table 3).

## 4 Discussion

This study found a strong association between musculoskeletal complaints and muscle stability deficits assessed by Adaptive Force in football players (OR = 54.0), with AF-Ratio discriminating clearly between players with and without complaints (*d* = −1.47, AUC = 0.93) while MVIC and H:Q ratio did not (AUC ≤ 0.44). Combined with the preliminary follow-up showing lower baseline AF-Ratio (*d* = −1.45, AUC = 0.86) and more stability deficits (*d* = 1.67, AUC = 0.90) in players with subsequent complaints, these findings suggest that adaptive holding capacity captures a clinically relevant dimension of neuromuscular function not assessed by traditional strength testing.

Region-specific analyses revealed a bidirectional pattern: when complaints were present, corresponding stability deficits were almost always detected (e.g., 100% of players with hamstring complaints had HAM deficits), whereas deficits frequently occurred without corresponding regional complaints. This suggests that muscle instability may be necessary but not sufficient for complaint development—functioning as a predisposing factor that increases vulnerability—while additional demands (physical strain) are required for clinical manifestation. Under load, stability deficits may leave joints inadequately stabilized: in unstable muscles, adaptive holding capacity reaches its limit at less than half of the muscle’s maximal force output (AF-Ratio ∼39%).[27] Once the muscle can no longer maintain its length against the rising load, the articulating structures are no longer kept in position and displace prematurely; passive tissues must absorb the load, being strained beyond their physiological tolerance—this can plausibly result in non-contact injuries or ‘overuse’ syndromes. AF was assessed in an unloaded position and as a snapshot; under weight-bearing conditions and with accumulated fatigue, stability deficits may be more pronounced— indicated by the high deficit rates in players with acute training-induced complaints. It is hypothesized that musculoskeletal complaints or non-contact injuries occur in related joints or muscles when the involved muscles are unstable under specific loaded conditions—the mechanism proposed as the functional instability syndrome (FIS).[18] Accordingly, it is not the external load per se (‘overload’) but the reduced individual load-bearing capacity that drives complaint development. The frequent unilateral occurrence of complaints despite symmetrical loading and the observed side concordance (82%, *d* = −0.77) further support this interpretation.

The AF-Ratio of hip abductors showed the strongest group difference between players with and without complaints (*d* = −1.64), with instability in 79% of symptomatic but none of the complaint-free players. Prospective data on ABD strength and injury risk are inconsistent.[8,11] Nevertheless, hip abduction and dynamic knee valgus were seen regularly during non-contact anterior cruciate ligament (ACL) injuries [17]—where abductors must adapt to external loads to control pelvic alignment, influencing the entire lower-extremity kinetic chain. Specifically during landing, abductor instability may increase knee valgus moments and thus ACL injury risk.[31] Impaired ABD holding capacity may therefore have implications beyond hip pathology. All five players with distal complaints showed ABD deficits (AF-Ratio: ∼30%) and the three with knee complaints had deficits in all muscles except MQF (most markedly in HFL and ADD), suggesting a prominent role of joint-stabilizing muscles, rather than primary force generators (MQF). These observations require cautious interpretation given the small subgroup sizes, but warrant further investigation.

Neither MVIC nor H:Q ratio discriminated players with vs. without musculoskeletal complaints or subsequent injuries—consistent with studies showing no or limited association between strength and complaints or injury risk.[5–11,32] MVIC, H:Q ratio and AF_max_ yielded AUC values below 0.5, i.e. no discrimination in the hypothesized direction (all CIs including 0.5). Even where a group-level association exists, it need not translate into discrimination between symptomatic and asymptomatic players—the property a screening test actually requires.[6] Bakken et al.[8] illustrate this: of 20 strength measures examined, only three were significantly associated with injury risk— in inconsistent directions, greater quadriceps strength accompanying higher risk but greater adductor strength lower risk—while even these showed poor discrimination (AUC = 0.45–0.56). These magnitudes are similar to the AUCs of strength-related parameters found here for the cross-sectional baseline (≤0.44) and for the preliminary follow-up (≤0.38). The agreement—with Bakken et al. at the level of discrimination, and with the broader literature showing no or limited strength–injury association—indicates that the low discrimination of strength parameters in the present data is a consistent finding and not a sample-specific or methodological artefact. Despite such unsatisfactory findings from decades of research, the focus has remained on strength-related parameters rather than on questioning whether force production is the appropriate construct.

The results suggest that the AF might offer an alternative: besides the clearly significant difference between complaint groups with large effect size, the AF-Ratio reached the highest AUC value of 0.93, which significantly differed in the direct comparison with the AUCs of MVIC, H:Q ratio and AF_max_ (*p* ≤ 0.036; Table 3). Notably, the two components of the AF-Ratio, derived from the same measurement, dissociated: AF_max_—the maximal force component—patterned with MVIC and the H:Q ratio, whereas AFiso_max_—the holding-capacity component—showed intermediate discrimination (AUC = 0.79) and did not differ significantly from AF-Ratio. This ordering localizes the discriminating feature in the adaptive holding capacity, not in maximal force output or the measurement procedure. Nor is it an artefact of the ratio form: the H:Q ratio did not discriminate and was uncorrelated with AF-Ratio (*ρ* = −0.18), supporting that they capture different aspects of neuromuscular function (within-muscle control quality vs. inter-muscular strength balance).

The same pattern emerged for the preliminary follow-up data: in contrast to MVIC and H:Q ratio, AF-Ratio clearly separated the complaint groups (*d* = −1.45, Table 2) and discriminated players who did and did not develop subsequent complaints (AUC = 0.863), with significance in the direct comparison with the AUCs of MVIC, H:Q and AF_max_ (*p* ≤ 0.028; Table 3). The number of stability deficits indicates that even a simple count of deficient muscles—without force quantification—captures much of the discriminative information for both baseline and follow-up complaints (AUCs ≥ 0.90).

Notably, all players with self-reported complaints were actively involved in preseason training and matches, in contrast to other studies and reviews on strength-related parameters, which mainly consider time-loss injuries. This indicates that the AF-Ratio might capture deficits before complaints are severe enough to cause any time loss. Moreover, most players with follow-up complaints also had baseline complaints. Together with the small subgroup size, this limits the interpretation as prediction of new injury, but aligns with the high re-injury rates in football,[5,11,12,33] where subclinical factors such as nociception or reduced functional capacity may persist even after return to full, asymptomatic activity and sustain injury vulnerability. As such disturbances cannot be reliably determined by conventional measures,[34] the AF assessment might help to detect them at a subclinical stage. This is supported by findings that the holding capacity immediately and significantly decreased in reaction to experimentally irritated proprioception in complaint-free, healthy individuals [18–20] and was found to be clearly impaired in conditions in which nociception is involved [18,24,35]—consistent with the present association between AF-Ratio and self-reported complaints. This indicates sensitivity to afferent and central influences not captured by conventional maximal strength.

The selective impairment of holding capacity—with preserved pushing strength—is consistent with the differentiation between holding (HIMA) and pushing/pulling (PIMA) isometric muscle action as neurophysiologically distinct motor control strategies, even under constant load.[36–38] The adaptive holding function—based on HIMA and particularly challenging the neuromuscular system due to the increasing external load—requires a continuous sensorimotor feedback loop with target-actual comparison of muscle length and tension, placing higher demands on neuromuscular control than the comparatively simpler motor drive underlying pushing/pulling actions. The sensorimotor feedback loop involves proprioceptive afferents and central structures including the sensorimotor cortex, thalamus, cerebellum, cingulate cortex, basal ganglia, amygdala, and the reticular formation,[18,39–46] which also receive inputs from stress-related axes, the autonomic nervous system, limbic structures and nociceptive afferents via the spinoreticular pathway.[46–48] The modulation of gamma motor neuron excitability via the reticulospinal tract may represent a key mechanism through which diverse inputs affect spindle sensitivity and thus adaptive holding capacity.[18] The convergence within the same neural structures provides a plausible neuroanatomical basis for the high vulnerability of the holding function; moreover, it may explain the immediate and reversible switching between stable and unstable states upon presentation of disruptive or supportive stimuli.[18–24,35] This sensitivity and immediate responsiveness to diverse modulatory influences enable targeted diagnostics: destabilizing factors can be identified individually, while stabilization upon supportive stimuli identifies potentially beneficial measures, guiding personalized interventions.[18,24,35] While conventional strength testing may inherently lack the sensitivity to detect such deficits, the AF concept offers a distinct alternative with high potential for diagnostic and therapy purposes.

The self-referenced AF-Ratio demonstrated consistent discriminative capacity across all muscle groups, both between stability categories (reported previously [27], unstable: 30–55%, stable: 99–100%) and between players with and without complaints, most prominently for ABD, HFL and HAM (Table 2). It is suggested therefore to reconsider the muscles prioritized in screening: joint-stabilizing muscles (ABD, HAM, HFL) rather than primary force generators (MQF). The muscle-independent property of AF-Ratio is consistent with pooled data across six studies, where AF-Ratio was robust across muscles, examiners, participant sex and experimental conditions,[18] supporting its potential for screening—capturing a dimension of neuromuscular function relevant to injury-prone movements. The strong association with complaints even in fully active players supports the hypothesis that muscle instability may represent a key predisposing factor for the development of musculoskeletal complaints and non-contact injuries—for which the FIS provides a mechanistic framework, integrating the neurophysiological basis outlined above.[18]

This also has implications for prevention, in that improvements in its effectiveness have been called for.[1] Based on the hypothesis underlying the FIS—instability results in higher vulnerability—AF-based prevention programs should focus on resolving muscular stability deficits. However, AF is likely not directly trainable through exercise, since instability appears driven not by physical conditioning deficits but by factors interfering with neuromuscular control (e.g., disturbed proprioception, nociception, emotional distress).[18–21,24] Addressing the underlying individual cause—using AF as a real-time diagnostic marker—may be a prerequisite for stability restoration after which targeted training could further improve holding capacity. Prevention programs incorporating both warrant investigation.

Overall, the findings suggest that AF-Ratio reflects a construct distinct from conventional strength parameters: it captures another dimension of muscle function, which includes the adaptive holding component that injury-prone situations demand, and showed both clear group-level differences and discrimination between players with and without complaints, indicating clinical relevance. AF assessment may identify neuromuscular impairments prior to manifestation as time-loss injuries, potentially also for return-to-sport decisions. The preliminary follow-up data provide tentative support. Given the marked separation of AF-Ratio—unlike the overlapping strength distributions—the continuous AF-Ratio can be translated into a dichotomous outcome of muscle stability: stable vs. unstable.[18,27] Bahr identifies exactly this step as a prerequisite for a test to be useful in clinical practice.[6] Whether AF-Ratio meets the standards of screening tests—a prospective, externally validated prediction of future injury [6]—remains to be established.

### Limitations

The sample was limited to male semi-professional players from a single team, restricting generalizability; the small complaint-free group (n = 4) limits statistical power and yields sparse cell counts; the odds ratio is therefore imprecise (wide CI) and its magnitude should be interpreted cautiously. The cross-sectional design precludes causal inference; follow-up data (n = 13) should be interpreted as hypothesis-generating.

Measurements during intensive preseason preparation may have transiently influenced deficit prevalence, although reflecting a practically relevant screening timepoint; this was addressed by classifying acute training-related complaints separately.

Complaints were self-reported via structured but non-validated questionnaires, with possible recall bias and communication barriers (multilingual sample). Examiners were aware of complaints, potentially introducing observer bias; however, analyses used algorithm-based classification derived from objective data, and similar force application was confirmed.

## 5 Conclusion

In semi-professional football players, musculoskeletal complaints were strongly associated with stability deficits, and the AF-Ratio distinguished affected from unaffected players where conventional strength parameters did not—both cross-sectionally and in preliminary follow-up. This suggests that adaptive holding capacity captures a clinically relevant dimension of neuromuscular function beyond maximal strength. The proposed concept of FIS provides a mechanistic framework for the complaint–instability association.

Based on current knowledge of AF, it is proposed to reconsider the construct assessed in research and practice: muscle stability—characterized by adaptive holding capacity—rather than maximal strength. The AF-Ratio—muscle-independent and self-referenced, thus requiring no normative reference values—captured deficits in players who remained fully active, before any time loss. AF assessment may therefore hold potential for screening and diagnostics—addressing a gap that decades of strength-based research have not closed.

The present findings warrant further investigation of whether stability assessment can prospectively identify players at risk, including for return-to-sport decisions, and whether stability-targeted interventions and prevention programs reduce complaints and injuries.

## Data availability statement

The datasets generated during and/or analyzed during the current study are available from the corresponding author on reasonable request.

## Acknowledgements

The authors thank the participating players and staff of the collaborating football club for their participation and support.

## Author contributions

All authors contributed to the study conception and design. Data collection was performed by LVS and FNB. Data analysis was performed by LVS and JU. LVS drafted the manuscript. All authors read and approved the final version.

## Competing Interests Statement

The authors declare that they have no competing interests.

## Funding

No funding was received for conducting this study.

## List of abbreviations

ABD: hip abductors
ACL: anterior cruciate ligament
ADD: hip adductors
AF: Adaptive Force
AFiso_max_: maximal isometric Adaptive Force
AF_max_: maximal Adaptive Force
AF-Ratio: ratio of AFiso_max_ to AF_max_
AUC: area under the curve
CI: confidence interval
FIS: functional instability syndrome
HAM: hamstrings (knee flexors)
HFL: hip flexors
HIMA: holding isometric muscle action
H:Q: ratio hamstrings-to-quadriceps ratio
ICC: intraclass correlation coefficient
LE: lower extremity
M: arithmetic mean
MQF: quadriceps femoris muscle (knee extensors)
MVIC: maximal voluntary isometric contraction
NPV: negative predictive value
OR: odds ratio
PIMA: pushing isometric muscle action
PPV: positive predictive value
ROC: receiver operating characteristic
SD: standard deviation
Sens: sensitivity
Spec: specificity

## Notes

### Competing Interest Statement

The authors have declared no competing interest.

### Summary of Updates

Title and Abstract were revised.

