## Supplementary Material for "Muscle stability deficits and musculoskeletal complaints in football players: Adaptive Force Ratio outperforms conventional strength parameters—a cross-sectional study with preliminary follow-up"

---

### Supplementary Methods

***Setting and testing procedure*** *(adapted from Schaefer et al. [1])*

Two experienced examiners conducted all tests (male and female; 30 and 14 years of test experience). MVIC and AF measurements were performed in five muscle groups bilaterally in standardized positions (Supplementary Figure S1): MQF (quadriceps femoris muscle; knee extension), HFL (hip flexors), ADD (hip adductors) and ABD (hip abductors) supine, HAM (hamstrings, knee flexion) prone.

Familiarization included device-free manual muscle tests to clinically assess the holding capacity (muscle stability) of six muscle groups bilaterally (HFL, ADD, ABD, HAM, pectoralis major and deltoideus muscles). Afterwards, MVIC was recorded twice per muscle and side (60 s rest): participants pushed maximally against the examiner's fixed manual resistance (force build-up 2–3 s, held ~1 s); for MQF the device was anchored to the table, as knee-extension force exceeded manual resistance. AF was then assessed three times per muscle and side (30 s rest) from identical positions: participants held the limb isometrically in the starting position while the examiner applied a progressively increasing force along a standardized

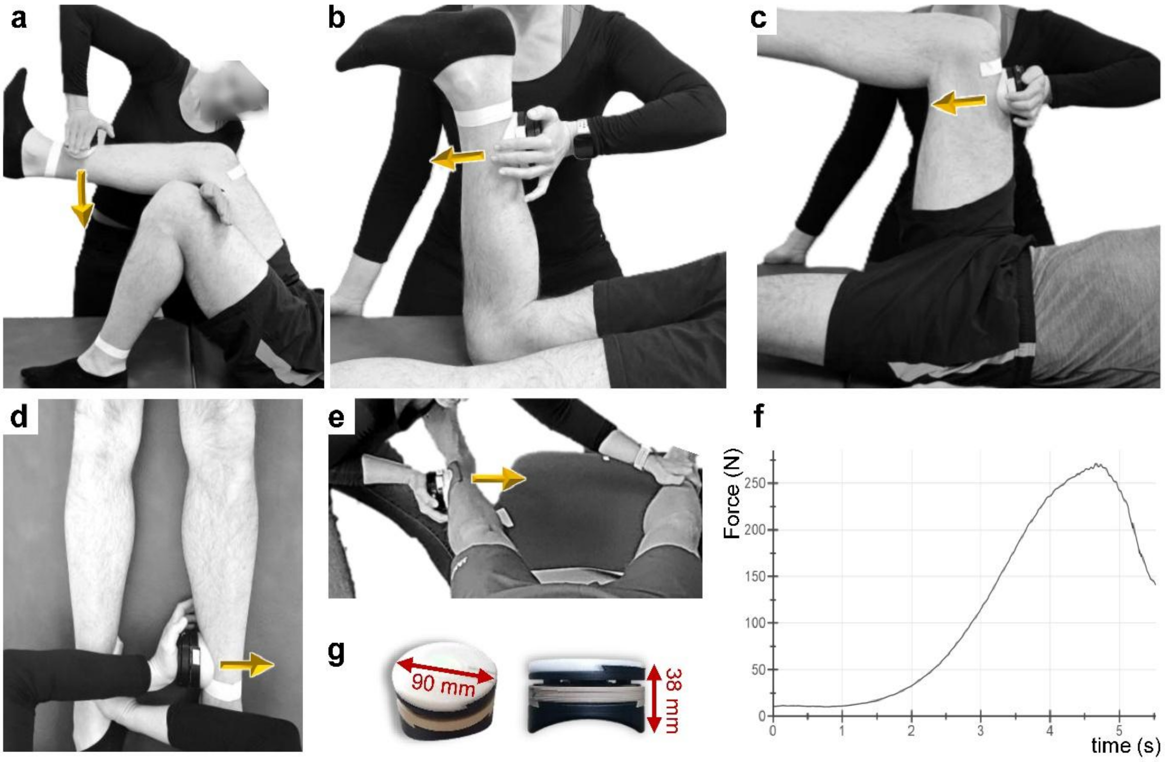

***Supplementary Figure S1. Starting positions of tested muscles and schematic force profile of AF trials.*** *The starting position and force vector applied by the tester corresponded to the plane of the main function of the muscle group being tested (**a**: MQF–knee extension, **b**: HAM–knee flexion, **c**: HFL–hip flexion, **d**: ADD–hip adduction, **e**: ABD–hip abduction). For AF measurements, the tester applied the increasing force (**f**: standardized force profile; max. force depends on test muscle and tester-participant-constellation) in the direction of muscle lengthening (arrow direction). The handheld device (**g**) was placed between the tester's palm and the participant's limb and measured force and position during the test. The same starting positions were used for MVIC tests, but the participant pushed against the fixed resistance provided by the tester (isometric activation in the direction of muscle shortening; arrow direction not shown). Reproduced from Schaefer et al.[1] (CC BY 4.0).*

profile (Supplementary Figure S1).[2,3] Trials were rated *stable* when the position was maintained, *unstable* when the limb involuntarily yielded (isometric-to-eccentric transition), and *borderline* otherwise.

#### *Data processing and parameter extraction*

Signals were processed in NI DIAdem 2019 and Python 3.12: interpolated to 1000 Hz (linear spline) and low-pass filtered (Butterworth, 20 Hz, $5^{th}$ order); absolute force parameters (N) were converted to torque (Nm; force × lever length). Five parameters were derived per muscle and side:

(1) MVIC: peak of two MVIC trials; higher of two trials was used for statistics.
(2) $AF_{max}$: peak force of AF trials [can arise during isometric (stable) or eccentric muscle action (unstable)]; arithmetic mean was used for statistics.
(3) $AFiso_{max}$: highest force maintained isometrically in the starting position, i.e. maximal holding capacity. $AFiso_{max}$ equals $AF_{max}$ when the starting position is held to peak force, and is lower when the limb yields earlier at submaximal force levels during the force increase (see Fig. 1 in main article). The identification of the onset of yielding is necessary to detect $AFiso_{max}$, reflecting the highest force under isometric conditions. This was detected from the gyroscope signal (gX): a zero-intercept least-squares linear fit ($gX = c \cdot F$) provided the slope c, and $AFiso_{max}$ was set to the force at which $c \cdot F(t)$ first exceeded a muscle-specific threshold (4°/s for HFL/MQF/HAM; 2°/s for ADD/ABD). This algorithmic estimate approximates the true $AFiso_{max}$—the force at the onset of muscle lengthening (break point). Lower thresholds for ADD and ABD accounted for their reduced test range of motion. Slight, spring-like yielding is a normal feature of muscular interaction and does not indicate instability or eccentric muscle action; at 4°/s the associated angular displacement remains minimal (<4° within the first second) and thus within the quasi-isometric range. Threshold robustness was confirmed by a sensitivity analysis of the $AFiso_{max}$-detection thresholds across five threshold sets[i] and two stability classification criteria to categorize stable and unstable muscles (clinical examiners' ratings and algorithm-based classification; see Supplementary Methods: Stability Classification), in which the AF-Ratio ($AFiso_{max}/AF_{max}$) consistently discriminated stable from unstable muscles [Cohen's $d$ = 4.30–10.01; all $p < 0.001$; examiners' ratings: $d$ = 4.8 (uniform 3°/s) to 10.01 (stricter); algorithm-based: $d$ = 4.3 (uniform 3°/s) to 9.0 (stricter)]; full details are reported in Schaefer et al. [1].
(4) AF-Ratio: $AFiso_{max}$ / $AF_{max}$ × 100 (muscle stability parameter)
(5) H:Q ratio: HAM MVIC / MQF MVIC × 100

[i] Reference: 4 and 2°/s, muscle-specific; stricter: 3 and 1.5 °/s, muscle-specific; more liberal: 5 and 2.5 °/s, muscle-specific; uniform 2°/s and uniform 3°/s, both across all muscles.

#### ***Stability classification***

Stability (stable, unstable, borderline) was classified automatically from the yielding parameter c · $AF_{max}$ (°/s) (quantifying the overall degree of muscular yielding; c from gX = c · F) using muscle-specific boundaries (Supplementary Table S1; full rules in Schaefer et al. [1]). Agreement with the examiners' independent clinical ratings was almost perfect (individual trials: overall three-category agreement (n = 645/685): 94.2%, κ = 0.89, quadratically-weighted κ = 0.97; two-category agreement (definite stable or unstable according to examiners' ratings, n = 619/627): 98.7%, weighted κ = 0.97). For statistical analyses, the stability classification was aggregated across the three trials to reveal one stability classification per muscle and side [n = 230; three-category agreement (215/230): 93.5%, κ = 0.88; weighted κ = 0.96; two-category agreement (199/202): 98.5%, no stable–unstable misclassifications]. The algorithm classification was used for statistics. Sensitivity analysis of stability-classification boundaries revealed similar results for stable vs. unstable classification across five classification methods and boundaries (Supplementary Figure S2; see Schaefer et al. [1] for details). Examiner-based discrimination consistently exceeded algorithm-based, supporting the algorithm as the more conservative, rater-independent criterion.

***Supplementary Table S1. Classification boundaries.***

<table>
<tr><th></th><th colspan="3">yielding parameter c · AFmax (°/s)</th><th></th><th>Euler-Parameters (°)</th></tr>
<tr><th>Muscle</th><th>Stable</th><th>Unstable</th><th>Borderline</th><th></th><th>Addition (borderline area)</th></tr>
<tr><td>HFL</td><td>< 4</td><td>> 6</td><td>4 – 6</td><td></td><td>—</td></tr>
<tr><td>MQF</td><td>< 3</td><td>> 6</td><td>3 – 6</td><td></td><td>—</td></tr>
<tr><td>HAM</td><td>< 3</td><td>> 5</td><td>3 – 5</td><td></td><td>—</td></tr>
<tr><td>ADD</td><td>< 1</td><td>> 3</td><td>1 – 3</td><td>1 – 3</td><td>stable: MM5020 < 6<br>unstable: MM5020 >8 AND MM9090 > 1<br>else: borderline</td></tr>
<tr><td rowspan="2">ABD</td><td rowspan="2">< 1</td><td rowspan="2">> 3.5</td><td rowspan="2">1 – 3.5</td><td>1 – 2.5</td><td>stable: MM8020 <5<br>else: borderline</td></tr>
<tr><td>2.5 – 3.5</td><td>unstable: MM8020 ≥6<br>else: borderline</td></tr>
</table>

MM = maximum monotonic (i.e., uninterrupted, without directional reversal) angular displacement in the yielding direction extracted from Euler angle signals within defined force ranges: MM5020 covers 50–20% $AF_{max}$ (broad range including the full yielding phase); MM8020 covers 80–20% $AF_{max}$; MM9090 covers 90–90% $AF_{max}$ (high-force region only)

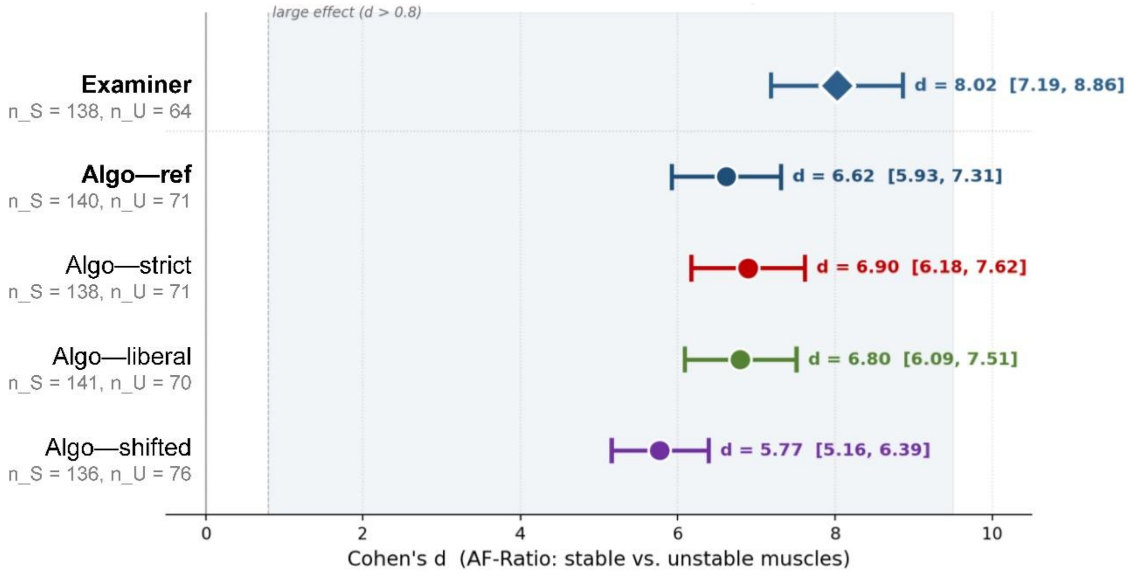

***Supplementary Figure S2. Sensitivity analysis of AF-Ratio group differences across classification methods and boundaries.***

The forest plot illustrates the effect sizes (Cohen's *d* with 95% confidence intervals) comparing AF-Ratio between stable and unstable muscles regarding the following different classification approaches: 'Examiner' (based on examiner's tactile-kinesthetic rating during AF assessment) and 'Algo' (algorithm-based on yielding coefficient $c \cdot AF_{max}$ (°/s), where c is the coefficient of the least-squares linear fit without intercept ($gX = c \cdot F$) of angular velocity (gX) and force (F) signals of AF trials). For algorithm-based classification, different boundaries for yielding coefficient were considered: ref = used for main analysis in the study, strict = lowered boundaries for stable/raised boundaries for unstable (broader borderline range), liberal = raised for stable/lowered for unstable (minimized borderline range), shifted = both boundaries lowered (borderline range constant). Concrete muscle-specific boundaries for stability classification (stable | unstable):

| | HFL | MQF | HAM | ADD | ABD |
|---|---|---|---|---|---|
| Reference | < 4 \| > 6 | < 3 \| > 6 | < 3 \| > 5 | < 1 \| > 3 | < 1 \| > 3.5 |
| Strict | < 3 \| > 7 | < 2 \| > 7 | < 2 \| > 6 | < 0.5 \| > 3.5 | < 0.5 \| > 4 |
| Liberal | < 4.5 \| > 5.5 | < 3.5 \| > 5.5 | < 3.5 \| > 4.5 | < 1.25 \| > 2.75 | < 1.25 \| > 3.25 |
| Shifted | < 3 \| > 5 | < 2 \| > 5 | < 2 \| > 4 | < 0.5 \| > 2.5 | < 0.5 \| > 3 |

n_S = number of stable muscles, n_U = number of unstable muscles *(adapted from Schaefer et al. [1])*

### Supplementary Data

#### *Rate of force development (RFD) as prerequisite for analyses*

As prerequisite for comparable force application across groups, the RFD of the phase of active force build-up before considerable yielding effects was calculated (linear regression on force signals from onset to 50% $AF_{max}$) and compared between stability categories. The non-significant differences (z-standardized, $p = 0.514$) confirmed the independence of force application rate (for more detailed information see Schaefer et al. [1]).

### Supplementary Table S2. Associations between complaint regions and stability deficits.

*Complaint → Deficit: proportion (%) of players with complaints in a given region who had stability deficits in the respective muscle group; n = number of players with complaints in that region.*
*Deficit → Complaint: proportion (%) of all players with a stability deficit in a given muscle group who reported complaints in that region. Total players with stability deficits per muscle group: HFL = 13, MQF = 5, HAM = 16, ADD = 9, ABD = 17. Players with complaints in multiple regions are counted in each applicable row.*

| Complaint region | n | Muscle | Complaint → Deficit | Deficit → Complaint |
|---|---|---|---|---|
| **Hip/groin/adductor** | 6 | **Any** | 5/6 (83%) | – |
| | | HFL | 4/6 (67%) | 4/13 (31%) |
| | | MQF | 1/6 (17%) | 1/5 (20%) |
| | | HAM | 2/6 (33%) | 2/16 (13%) |
| | | ADD | 3/6 (50%) | 3/9 (33%) |
| | | ABD | 4/6 (67%) | 4/17 (24%) |
| **Hamstrings** | 4 | **Any** | 4/4 (100%) | – |
| | | HFL | 2/4 (50%) | 2/13 (15%) |
| | | MQF | 0/4 (0%) | 0/5 (0%) |
| | | HAM | 4/4 (100%) | 4/16 (25%) |
| | | ADD | 2/4 (50%) | 2/9 (22%) |
| | | ABD | 4/4 (100%) | 4/17 (24%) |
| **Knee** | 3 | **Any** | 3/3 (100%) | – |
| | | HFL | 3/3 (100%) | 3/13 (23%) |
| | | MQF | 0/3 (0%) | 0/5 (0%) |
| | | HAM | 2/3 (67%) | 2/16 (13%) |
| | | ADD | 3/3 (100%) | 3/9 (33%) |
| | | ABD | 2/3 (67%) | 2/17 (12%) |
| **Distal lower extremity** | 5 | **Any** | 5/5 (100%) | – |
| | | HFL | 4/5 (80%) | 4/13 (31%) |
| | | MQF | 1/5 (20%) | 1/5 (20%) |
| | | HAM | 4/5 (80%) | 4/16 (25%) |
| | | ADD | 3/5 (60%) | 3/9 (33%) |
| | | ABD | 5/5 (100%) | 5/17 (29%) |
| **Load-related/generalized** | 6 | **Any** | 6/6 (100%) | – |
| | | HFL | 4/6 (67%) | 4/13 (31%) |
| | | MQF | 4/6 (67%) | 4/5 (80%) |
| | | HAM | 5/6 (83%) | 5/16 (31%) |
| | | ADD | 3/6 (50%) | 3/9 (33%) |
| | | ABD | 6/6 (100%) | 6/17 (35%) |
| **Upper extremity** | 2 | **Any** | 2/2 (100%) | – |
| | | HFL | 2/2 (100%) | 2/13 (15%) |
| | | MQF | 1/2 (50%) | 1/5 (20%) |
| | | HAM | 2/2 (100%) | 2/16 (13%) |
| | | ADD | 1/2 (50%) | 1/9 (11%) |
| | | ABD | 2/2 (100%) | 2/17 (12%) |

*ABD = hip abductors; ADD = hip adductors; HAM = knee flexors (hamstrings); HFL = hip flexors; MQF = knee extensors (quadriceps femoris). Any = stability deficit present in at least one of the five muscle groups (pooled). Load-related/generalized = acute muscle soreness or pain related to training load.*
